# Diverging Patterns of Cognitive Decline by Sex and Race-Ethnicity in Seriously Ill Older Americans

**DOI:** 10.1101/2024.06.27.24309609

**Authors:** Iván Mejía-Guevara, V.S. Periyakoil

## Abstract

**Objectives:** Differences in Cognitive decline are common in older adults in the last years of life, but differences across sex and race-ethnicity are poorly understood. This study investigated if sex and/or race-ethnicity moderated changes in cognitive function in older adults in the last years of life.

**Methods:** Data from the Health and Retirement Study (HRS) from 1993 to 2016 were used to analyze imputed cognition summary scores for total word recall and mental status of older adults aged 60-99. Loss of cognitive function was estimated using a multilevel mixed-effects model and accelerated cognitive decline was approximated by incorporating a change-point model using a restricted sample of decedent respondents who died aged 65-99.

**Results:** Notable disparities were seen in the rates of cognitive decline across sex and race-ethnic groups in the last years of life. Women consistently scored lower than men in word recall but higher in mental status, regardless of race-ethnicity. Non-Hispanic White respondents, men and women, consistently outperformed Hispanic and Black respondents in word recall tasks and mental status.

**Conclusions:** Our study shows that sex and race-ethnicity moderate cognitive decline in older adults during the last years of life. Older adults from underserved communities are at higher risk of cognitive decline. Our study could inform clinical practice and policy focused on mitigating the adverse impact of cognitive decline experienced by marginalized populations of older adults in the last years of life.

## Introduction

**The impact of aging on cognition is clear, but sex differences in cognitive changes over time in older adults are unclear** (Miller & Halpern, 2014). Older women may have higher cognitive reserve but faster cognitive decline than older men (Levine et al., 2021). Older women also perform better than men in most standardized cognitive tests, except for spatial ability (McCarrey, 2016; Miller & Halpern, 2014). In addition, mixed results have been found regarding cognitive decline by race among older adults without dementia, some reporting no race differences in cognitive decline (Castora-Binkley et al., 2015; Karlamangla et al., 2009). Race and ethnic differences in cognitive decline are known, with Black Americans and Hispanic or Latino individuals showing a higher incidence of dementia than non-Hispanic White (NHW) populations (Mehta & Yeo, 2017). A recent study has identified significant race and ethnic differences in mild cognitive impairment and dementia, with Hispanic or Latino groups showing higher risk than NHW and Black or African American groups (Wright et al., 2021), although cognitive impairment data on Hispanic or Latino individuals are more limited.

**Little is known about sex and race-ethnic combined differences in seriously ill older adults in the last years of life.** There is a well-established survival advantage of females relative to males, and mortality disparities across racial and ethnic groups in old age (Crimmins, 2018). Knowledge about cognitive patterns in the last years of life will serve as a critical piece of data as we seek to mitigate late-life disparities in seriously ill older adults.

Dementia in older adults strongly predicts death, there is a competing risk of death that may affect the accuracy of estimates of cognitive function and decline (Hayes-Larson et al., 2020). Selective survival has been described as a phenomenon wherein individuals with specific characteristics disproportionally survive to old age while those characteristics are also associated with dementia risk (Hayes-Larson et al., 2020; Shaw et al., 2021a). It is also known that cognition declines in the last years of life (Riegel & Riegel, 1972; Wilson et al., 2003). In this study, we sought to explore the relationships between cognitive decline and its impact (if any) on mean age trajectories of cognitive function by sex and race-ethnicity using a national sample of older survivors and decedents.

## Method

### Study design and participants

The Health and Retirement Study (HRS) is a comprehensive, nationally representative longitudinal survey encompassing over 43,000 Americans aged 50 and older (HRS, 2022). Its primary objective is to monitor changes in the health, wealth, social structure, and functional abilities of its participants as they age.

For this study, we retrieved information on demographics, cognitive function scores, and mortality status for respondents aged 50 and older from eleven survey years (biennially 1996-2016) of the latest version of the RAND HRS Longitudinal File 2020 (v1, March 2023), which contains information from Core and Exit Interviews of the HRS, and revised cognitive scores during this period (Health and Retirement Study, 2023; RAND HRS Longitudinal File 2020 (V1), 2023). Our analysis focused on alive and decedent respondents aged 65-99, with decedents dying between the ages of 65-99 years in three self-reported racial and ethnic groups: Hispanic or Latino, non-Hispanic White, and non-Hispanic Black or African American (hereafter Hispanic, White, and Black, respectively). Mortality status was ascertained based on an interview status variable indicating whether the respondent had passed away between the last and the current interview or before a previous wave (Health and Retirement Study, 2023). Since our analysis focused on respondents aged 65-99 at the time of measurement, disregarding observations from individuals younger or older than this age range, the total number of respondents between 1996 and 2016 was 113,139 participants, 46,844 survivors, and 66,295 deceased, after these exclusions (**Table 1**).

**Table 1.** Sample size and descriptive values for survivors’ and decedents’ cognitive scores, exposure, and risk factors.

| Variable | Total | Survivor | Decedent* |
| --- | --- | --- | --- |
| No. respondents by wave |  |  |  |
| Total 1996-2016 | **113,139** | **46,844** | **66,295** |
| 1996 | 8,049 | 731 | 7,318 |
| 1998 | 10,425 | 1616 | 8,809 |
| 2000 | 10,379 | 1999 | 8,380 |
| 2002 | 10,581 | 2743 | 7,838 |
| 2004 | 10,768 | 3464 | 7,304 |
| 2006 | 11,065 | 4316 | 6,749 |
| 2008 | 11,025 | 5067 | 5,958 |
| 2010 | 10,635 | 5749 | 4,886 |
| 2012 | 10,439 | 6366 | 4,073 |
| 2014 | 10,077 | 7010 | 3,067 |
| 2016 | 9,696 | 7783 | 1,913 |
| Age at first cognition report,<br>median (IQR) |  |  |  |
| Total word recall | 67 (65-73) | 66 (65-67) | 71 (66-75) |
| Total mental status | 67 (65-73) | 65 (66-67) | 71 (66-75) |
| Weighted mean cognitive score<br>(SD) across all Cognitive scores |  |  |  |
| Total word recall | 9.1 (3.6) | 10.1 (3.2) | 8.2 (3.7) |
| Total mental status | 12.7 (2.5) | 13.1 (2.1) | 12.3 (2.8) |
| No. cognition reports, median<br>(IQR) |  |  |  |
| Total word recall | 6 (4-8) | 7 (4-9) | 6 (3-8) |
| Total mental status | 6 (4-8) | 7 (4-9) | 6 (3-8) |
| **Non-modifiable factors** |  |  |  |
| No. observation in each age category at baseline (weighted %) |  |  |  |
| 65-69 | 1469 (5.9) | 369 (30.2) | 1100 (4.4) |
| 70-74 | 2020 (24.5) | 200 (32.6) | 1820 (24.6) |
| 75-79 | 2115 (32.8) | 111 (25.9) | 2004 (33.9) |
| 80-84 | 1471 (21.4) | 40 (9) | 1431 (21.9) |
| 85-89 | 814 (10.9) | 9 (2.1) | 805 (11.2) |
| 90-99 | 337 (4.5) | 2 (0.3) | 335 (3.9) |
| Age across all cognition reports, median (IQR) |  |  |  |
| Total word recall | 74 (69-80) | 71 (68-76) | 76 (71-82) |
| Total mental status | 74 (69-80) | 71 (68-76) | 76 (71-82) |
| Sex, N at baseline (weighted %) |  |  |  |
| Women | 4585 (57.8) | 446 (66.2) | 4139 (56.6) |
| Men | 3464 (42.2) | 285 (33.8) | 3179 (43.4) |
| Race/ethnicity, N at baseline (weighted %) |  |  |  |
| Hispanic or Latino | 496 (4) | 51 (5.7) | 445 (3.9) |
| non-Hispanic White | 6504 (88.2) | 608 (87.2) | 5896 (88.3) |
| non-Hispanic Black or African American | 1046 (7.8) | 71 (7) | 975 (7.8) |
| Education, N (weighted %) |  |  |  |
| Less than HS or GED | 3445 (41.2) | 226 (32.1) | 3219 (41.8) |
| HS graduate | 2361 (29.9) | 220 (30.2) | 2141 (30.2) |
| Some college | 1256 (16.2) | 151 (20.9) | 1105 (15.8) |
| College and above | 986 (12.7) | 134 (17.5) | 852 (12.2) |
| APOE-4, mean across all waves (95% CI) | 23.7 (20.6 - 27.1) | 17.6 (11.7 - 25.7) | 25.1 (21.5 - 29) |
| **Modifiable factors, weighted percentage across all measures (95% CI)** |  |  |  |
| Current cigarette smoking | 9.3 (8.6 - 10.1) | 7.7 (7 - 8.6) | 10.9 (10 - 11.8) |
| Alcohol drinks, No./wk. |  |  |  |
| None | 67.8 (66.3 - 69.4) | 61.4 (59.4 - 63.3) | 73.4 (71.9 - 74.9) |
| 1-6 | 19.9 (18.8 - 21.1) | 25.1 (23.6 - 26.6) | 15.4 (14.4 - 16.5) |
| 7-13 | 6.3 (5.9 - 6.7) | 7.1 (6.6 - 7.6) | 5.6 (5.1 - 6.1) |
| >=14 | 6 (5.6 - 6.4) | 6.5 (5.9 - 7.1) | 5.6 (5.1 - 6.1) |
| Body mass index |  |  |  |
| < 18.5 | 2.3 (2.1 - 2.5) | 1.1 (0.9 - 1.3) | 3.3 (2.9 - 3.6) |
| 18.5-25 | 34.1 (33 - 35.2) | 28.5 (27.2 - 29.9) | 38.9 (37.6 - 40.1) |
| 25-30 | 37.8 (37.2 - 38.5) | 40.2 (39 - 41.4) | 35.8 (35 - 36.6) |
| >=30 | 25.8 (24.8 - 26.8) | 30.1 (28.9 - 31.4) | 22.1 (21.1 - 23.1) |
| High blood pressure | 4.2 (4.1 - 4.4) | 4 (3.8 - 4.2) | 4.4 (4.3 - 4.6) |
| Diabetes | 2.5 (2.4 - 2.7) | 2.6 (2.4 - 2.8) | 2.5 (2.4 - 2.7) |
| Hearth disease | 4.2 (4 - 4.3) | 3.3 (3.1 - 3.4) | 5 (4.8 - 5.2) |
| Stroke | 2.1 (2.1 - 2.2) | 1.2 (1.1 - 1.3) | 3 (2.9 - 3.2) |
| Cancer | 2.7 (2.6 - 2.9) | 2 (1.8 - 2.2) | 3.4 (3.2 - 3.6) |
| Depression (CES-D score) | 1.4 (1.4 - 1.4) | 1.1 (1.1 - 1.2) | 1.7 (1.6 - 1.7) |

\* Values and estimates for decedents correspond to observations provided when they were alive in the specific period.

### Ethics Review

This study relied on publicly available data and no review process was needed.

### Outcomes

We considered two primary outcomes in this paper: (1) a total word summary score (TR20, range 0-20), calculated as the sum of immediate and delayed word recall scores; and (2) a mental status summary score (MSTOT, range 0-15), derived from a series of tests that evaluated knowledge, that included serial 7’s, backwards counting, date naming, object naming, and naming the President and Vice President of the United States (see Appendix for details of the cognitive tests). All these scores were retrieved from the HRS imputations of cognitive functioning (McCammon et al., 2022).

### Mortality and cognitive decline in the last phase of life

To investigate the relation of death to changes in cognitive function, we included mortality status to account for the impact of mortality on decedents’ outcomes to construct age trajectories of cognition. Further, our goal was to investigate the influence (if any) of approaching mortality on cognition (Wilson et al., 2003). Specifically, we built on previous research to estimate the onset time of cognitive decline in the last phase of life relative to the time of death and the rates of cognitive decline and cognitive scores during two distinct phases, a phase of cognitive decline and a phase of accelerated cognitive decline, demarcated by this initial timing (Sliwinski et al., 2006; Wilson et al., 2018, 2020). For this latter analysis, we restricted the analytic sample to all decedent respondents who died between 1996 and 2016, aged between 65 and 99. Additionally, we stratified the sample by sex and race-ethnicity to examine potential sub-group variations of cognitive decline before death. We used the term sex here because in HRS, sex and gender are used interchangeable (Hanes & Clouston, 2021).

### Non-modifiable and modifiable risk factors

We account for non-modifiable risk factors, including sex, age, race-ethnicity, and years of school, as self-reported by participants (Bloomberg et al., 2021). We also considered the APOE isoform, directly genotyped when available and imputed otherwise, which has three major isoforms (□2, □3, and □4), determined by two SNPs, rs7412 and rs429358. We inferred the APOE isoform based on the combination of these major isoforms and categorized it into six pairs: □2/□2, □2/□3, □2/□4, □3/□3, □3/□4, and □4/□4 (Faul et al., 2021). We recoded the APOE isoform as a binary factor, where respondents with one or two copies of the 4 allele (□2/□4, □3/□4, □4/□4) were assigned a value of one, while those without the 4 allele (□2/□2, □2/□3, □3/□3) were assigned a value of zero. We referred to this resulting variable as APOE-4, which has been significantly associated with Alzheimer’s disease and dementia in various studies (Safieh et al., 2019; Tang et al., 1998).

Modifiable factors, such as alcohol consumption (measured by the number of drinks per week), current smoking status, and body mass index (calculated as weight in kilograms divided by height in meters squared), and chronic diseases (diabetes, high blood pressure, heart disease, and stroke, using indicators that stipulated whether a doctor had ever diagnosed the respondent with any of these conditions) were included in the analysis (Health and Retirement Study, 2022; RAND HRS Longitudinal File 2018 (V2), 2022).

### Statistical Analysis

To assess changes in cognitive function/decline across ages before death, we utilized a mixed-effects change point model (see Appendix A in the Supplementary Material for a detailed analytic description of the model) (Hall et al., 2000; Karr et al., 2018; Sliwinski et al., 2006; Walter et al., 2016). This model combines elements from mixed-effects and change-point models to capture both fixed and random effects effectively. It aims to identify potential shifts in cognitive function at two different stages relative to the end of life, including the phase when cognitive decline accelerates before death.

#### Mixed-Effect Model: age trajectories of cognitive decline

To build age trajectories of cognition, we build up this model hierarchically, first estimating an intercept and age slope of cognition for each individual and then estimating the determinants of the person-specific intercepts and age coefficients (Karr et al., 2018; Walter et al., 2016). In our first model (equation 1 in Appendix A), we consider all survivors and decedents, ignoring the potential impact of accelerated cognitive decline before death; that is, we assume cognitive decline only depends on the chronological age, regardless of the time of death, where the onset time of the last phase relative to the time of death (represented by τ along the article) is equal to zero (see equation 1 in Appendix A with τ=0). In this model, we accounted for the age of each respondent over time, and for all covariates described in the previous section (except sex and race-ethnicity), including person-level variables (education, APOE) and those that can change with age for every individual (risk factors and health conditions). We included death status as an explanatory factor to account for the impact of mortality on the outcome of decedents in our model. The influence of sex and race-ethnicity is modelled by specifying individual intercepts and age slopes in the model (see equations 2-3 in Appendix A).

#### Change Point Model: cognitive decline acceleration in the last phase of life

To evaluate the potential acceleration of cognitive decline preceding death, we restricted the sample to all decedent respondents, and we relied on the change point model specification of equation 1 (Appendix A), which provides an adequate index of time to death, as opposed to traditional approaches that use the chronological age or time from birth (Sliwinski et al., 2006; Wilson et al., 2020). Therefore, the first model (see equation 1 in Appendix A, with r :;t:0) represents two change processes: a decline phase and a phase of accelerated variation. The decline phase uses chronological age to index normative age-related processes. In contrast, the accelerated phase represents two effects, one capturing normative age effects and those related to the progressive pathologies that lead to death (Sliwinski et al., 2006). To estimate the influence of age, sex, and race-ethnic differences in cognitive function before death during the decline and accelerated phases, we continued using the models specified in equations 1-3 (Appendix A).

Further details about the statistical model and estimation process are in Appendix A. All analyses were performed using ®Stata/SE 16.1 for Mac (StataCorp, College Station, TX).

#### Sensitivity Analysis

We conducted two sensitivity analyses. First, we replicated the statistical analysis by incorporating respondents with the APOE-4 variant into the change point model (equations 1-3) to assess differences in the rate of cognitive decline during the early and accelerated phases. Second, to evaluate the robustness and generalizability of our model, we employed a 10-fold cross-validation approach. This involved replicating the original sex and race-ethnic stratified analyses using the change point model to estimate rates of cognition change during the early and accelerated phases. This method relies on out-of-sample predictions to assess the consistency of coverage across validation sets, providing evidence of the stability and generalizability of the model. Further details of this procedure are provided in Appendix A of the Supplementary Material.

## RESULTS

### Descriptive data

We collected data on total word recall and mental status from the RAND-HRS biennial dataset from 1996 to 2016 (waves 3-13) after excluding waves 1 and 2, due to a different approach used in constructing the summary scores, and waves 14 and 15 due to the lack of updated reports on these specific indicators, which are under current revision in the latest version of RAND-HRS (Health and Retirement Study, 2023). About 58% of respondents were women, 88.2% of total respondents were White, 7.8% were Black, and 4.0% were Hispanic (**Table 1**).

During the study period, the median number of cognitive reports per respondent was 6 (interquartile range (IQR): 4-8). The mean word recall score across all waves was 10.1 (standard deviation [SD] = 3.2) for survivors and 8.2 (SD = 3.7) for decedents, while the mean mental status score for survivors was 13.1 (SD = 2.1) and 12.3 (SD = 2.8) for decedents. The median age across all mental status reports was 71 (IQR: 68-76) for survivors and 76 (71-82) for decedents (**Table 1**).

Regarding risk factors, decedents were more likely than survivors to be less educated, and exhibit higher prevalence of modifiable risk factors, including smoking, higher blood pressure, heart disease, stroke, cancer, and depression (**Table 1**)

### Functional cognitive decline: age trajectories by sex and race-ethnicity

Figure 1 depicts the relationship between age and cognitive function, with higher scores indicating better performance. The results show that cognitive function significantly declines with age. Irrespective of race and ethnicity, women outperformed men in word recall at all ages, while men scored higher than women in total mental status. Additionally, there were significant differences in cognition across ages among different racial and ethnic groups. Regardless of sex, White respondents exhibited higher word recall and mental status than Hispanic and Black respondents of all ages. Although Hispanic respondents had higher scores than Black respondents, the gap between the scores of White and Hispanic groups was more significant than those between Hispanic and Black respondents, even for men’s total mental status, where the gap between Hispanic and Black respondents was the highest (Figure 1). Figure S1 (in Appendix B in the Online Supplementary file) displays the associations by sex and race-ethnicity separately, confirming that women’s total word recall was higher than men’s, while men’s mental status was higher than women’s at all ages. In addition, it also confirms racial-ethnic differences in both scores. All these results were obtained after controlling for explanatory factors, the risk of mortality, and the APOE-4 variant.

**Figure 1.**
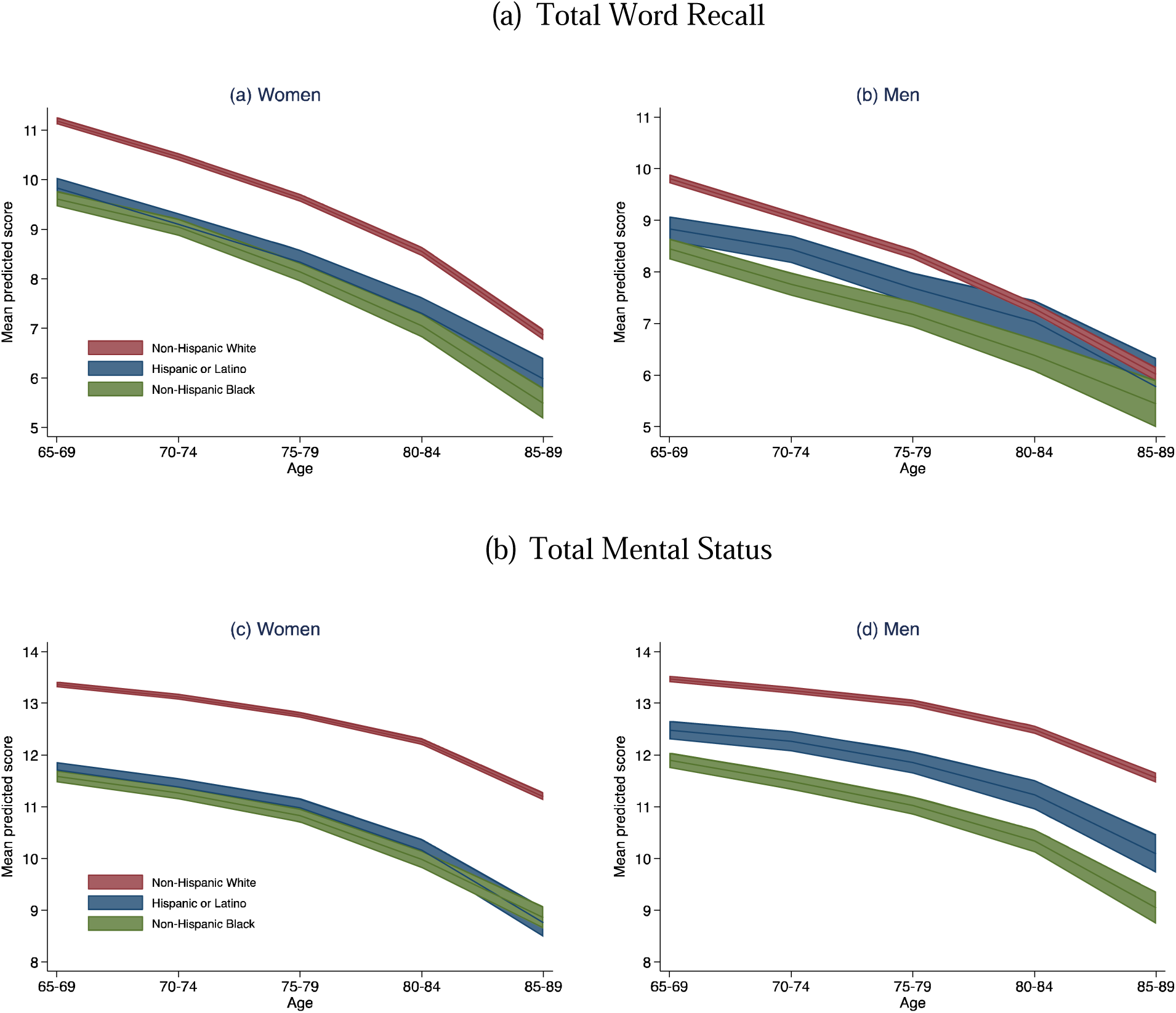
Predicted summary scores for the total word recall, and total mental status by age, sex, and race-ethnicity. Note: Age trajectories of cognition were estimated using the model specification of equations 1-3 in Appendix A, but with T=0.

### Early and accelerated stages of cognitive decline: sex and race-ethnic differences

Since the HRS does not provide the dementia status of all respondents in the sample, which could significantly bias the results of cognitive function/decline, we excluded respondents with the APOE-4 variant, as this gene variant has been highly associated with dementia in the literature. Table 2 presents the results of these models, including estimated rates during the early and accelerated phases of cognitive decline in stratified models by sex and race-ethnicity of deceased respondents. The table presents estimates of the onset time of the accelerated phase relative to the time of death (r) for individuals without the APOE-4 variant. On average, the onset time of accelerated decline for word recall was 10 years for all race-ethnic groups, except for Hispanic women and White men whose rate was 9 years. During the early phase, the cognitive decline rates were significantly negative for both scores and all race-ethnic groups. However, these rates accelerated for all groups after the onset or during the accelerated phase, resulting in a total negative effect. For instance, White and Black women experienced around a 20% decline in total word recall during the early phase, which increased to 36% decline for White women and 31% for Black women during the accelerated phase. In relative terms, the annual rate of accelerated decline (ARAD = accelerated decline rate/r) during the latter phase for White women was 3.6%, the highest rate observed among all race-ethnic and gender groups. Regarding mental status in women, Black women had the highest ARAD at 3.0%, with an additional cognitive decline of 27% attributable to accelerated phase, while the respective ARADs for White and Hispanic women were 2.8% and 2.2%. When combining women’s and men’s decline rates, White respondents exhibited more significant rates of cognitive decline during the accelerated phase for word recall, but closely followed by Black respondents in the case of mental status (Table 2).

**Table 2.** Point estimates of the onset time of the accelerated and rates of early and accelerated cognitive decline across sex and race-ethnic groups.

| Cognition Score | Phase 1: Early cognitive decline phase | Phase 2: Accelerated cognitive decline phase | Mean time in years of accelerated cognitive decline (Phase 2) | Annual Rate of Accelerated Decline (ARAD) |
| --- | --- | --- | --- | --- |
| **Total Word Recall** |  |  |  |  |
| Hispanic women | -0.19** (-0.22, -0.17) | -0.26** (-0.31, -0.22) | 9 | -2.9 |
| Hispanic men | -0.14** (-0.18, -0.11) | -0.24** (-0.28, -0.20) | 10 | -2.4 |
| White women | -0.20** (-0.20, -0.19) | -0.36** (-0.37, -0.35) | 10 | -3.6 |
| White men | -0.16** (-0.17, -0.15) | -0.30** (-0.31, -0.29) | 9 | -3.3 |
| Black women | -0.19** (-0.21, -0.17) | -0.31** (-0.34, -0.28) | 10 | -3.1 |
| Black men | -0.15** (-0.17, -0.13) | -0.23** (-0.26, -0.2) | 10 | -2.3 |
| **Total Mental Status** |  |  |  |  |
| Hispanic women | -0.14** (-0.16, -0.11) | -0.22** (-0.26, -0.19) | 10 | -2.2 |
| Hispanic men | -0.09** (-0.12, -0.07) | -0.21** (-0.25, -0.18) | 9 | -2.4 |
| White women | -0.08** (-0.09, -0.08) | -0.26** (-0.26, -0.25) | 9 | -2.8 |
| White men | -0.06** (-0.07, -0.05) | -0.20** (-0.21, -0.19) | 9 | -2.2 |
| Black women | -0.12** (-0.14, -0.11) | -0.27** (-0.30, -0.25) | 9 | -3.0 |
| Black men | -0.09** (-0.12, -0.07) | -0.22** (-0.25, -0.19) | 10 | -2.2 |

Note: For this analysis, we excluded respondents with the APOE-4 gene variant. The onset time refers to the accelerated cognitive decline phase relative to the time of death ( $\tau$ ). The ARAD is estimated as the rate of accelerated decline dividend by the onset time of accelerated decline. Levels of significance: \*( $p < 0.05$ ), \*\*( $p < 0.01$ ).

Figure 2 illustrates the predicted mean word recall and mental status scores around age 75 during the early and accelerated phases. For both scores and two phases, White women’s and men’s scores outperformed their Hispanic and Black counterparts. In addition, word recall remained high for women than men for all race-ethnic groups during the early and accelerated phases, while men outperformed women in total mental status during both phases as well. All these differences were significant for white groups while less precise for minoritized groups in some cases, perhaps due to the much larger sample of white respondents in the HRS study.

**Figure 2.**
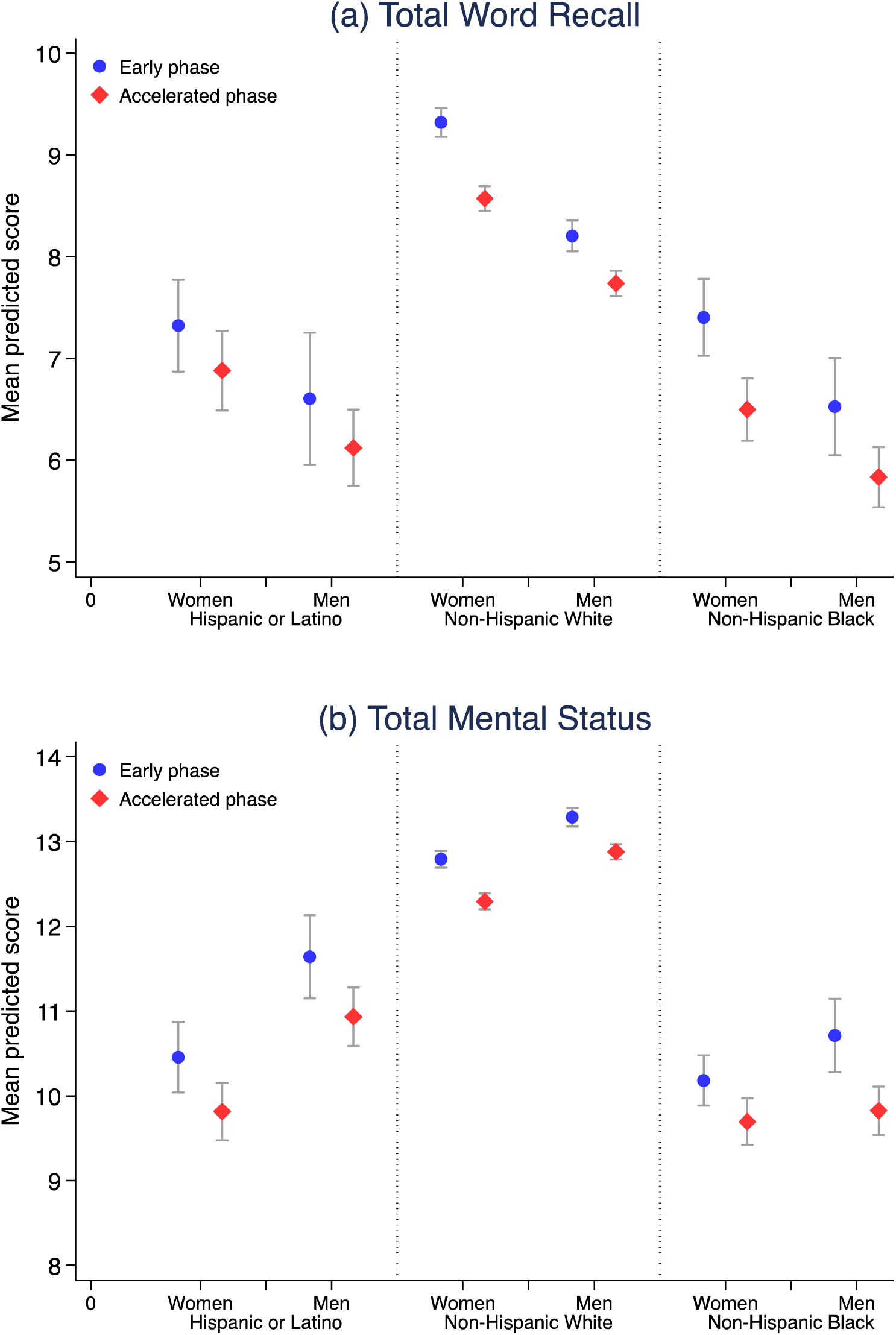
Mean predicted word and mental scores at age 75 from HRS decedents by sex and race-ethnicity. Note: Values predicted cognition were estimated using the change-point model specification from equations 1-3 in Appendix A, and excluding respondents with the APOE-4 gene variant.

### Sensitivity analysis

For the sensitivity analysis, we included deceased participants with the APOE-4 variant and replicated the results from Table 2 and Figure 2 using validation samples. Upon inclusion of respondents with the APOE-4 variant, certain groups exhibited minor variations in the τ value and rates of early and accelerated cognitive decline, although to a lesser extent regarding mental status (Table S1). This outcome was anticipated as we intentionally included individuals with a higher likelihood of cognitive impairment, given the strong relationship between APOE-4 and dementia. However, the predicted values for mental scores at age 75, stratified by race-ethnicity and gender groups (Figure S2), remained largely unchanged compared to those presented in Figure 2. The results were also consistent at all ages, with those obtained in the original analysis, as reported in Table S2 in Appendix B of the Supplementary file.

The sensitivity analysis conducted after applying 10-fold cross-validation to replicate the results shown in Figure 1 consistently confirmed the differences in cognitive function and rates of decline among different race-ethnic and gender groups, as initially predicted in the original analysis. These findings are depicted in Figure S3 and Table S2.

## DISCUSSION

This study has three salient findings. First, it shows sex and race-ethnic significant differences in cognitive function among older U.S. individuals. Irrespective of race-ethnicity, women perform better in word recall, while men exhibit higher mental status scores. Regardless of sex, Black respondents had lower performance in both word recall and mental status than Hispanic and White respondents. Second, we identified minor sex and race-ethnic differences in the estimated onset time of accelerated cognitive decline before death, but more significant differences in the rates of cognitive decline during both the early and accelerated phases. Third, we found compelling evidence that sex differences persist until the end of life, with women outperforming men in word recall and men surpassing women in mental status in both the early and accelerated phases of cognitive decline.

Our findings align with previous research indicating that women have lower performance levels in active skills like spatial and numerical ability, whereas men have lower levels in passive activities, such as verbal meaning, word fluency, and inductive reasoning (B. Bosworth, 1999; Levine et al., 2021; McCarrey, 2016). Our findings were obtained after controlling for non-modifiable and modifiable risk factors, including educational level, race, vascular risk factors, health conditions associated with increased risk of cognitive decline and dementia, mortality status, and APOE-4 gene variants (Bloomberg et al., 2021; Levine et al., 2020, 2021; Miller & Halpern, 2014; Takeda et al., 2020). Additionally, these findings also align with previous research indicating that African American and Hispanic residents are more likely to experience cognitive decline compared to non-Hispanic White residents, highlighting the presence of significant decline differences across race-ethnic groups. Black and Hispanic groups bear a higher burden of chronic conditions and adverse social determinants of health, which predict worse cognitive outcomes, including Subjective Cognitive Decline (SCD), SCD-associated functional limitations, cognitive function, or dementia incidence (Díaz-Venegas et al., 2016; Gupta, 2021; Kornblith et al., 2022; Wooten, 2023). Our results further contribute to this evidence by showing that racial-ethnic disparities persist irrespective of sex, although the differences in mental status scores between Hispanic and Black women are significantly less pronounced, with scores consistently lower than those of NHW women across all age groups.

Our results also confirm previous findings that cognitive decline accelerates in the last years of life (MacDonald et al., 2011; Riegel & Riegel, 1972; Sliwinski et al., 2006; Wilson et al., 2003, 2020). The decline rates are more significant among women maybe because of their higher longevity and prevalence of Alzheimer’s disease than men (Karlamangla et al., 2009; Levine et al., 2021; Shaw et al., 2021b; Alzheimer’s Association, 2023). Differences across race-ethnic groups varied depending on sex, outcome, and phase, but irrespective of sex, NHW respondents exhibited more significant rates of cognitive decline in the last years of life (Karlamangla et al., 2009; Weuve et al., 2018). Finally, the sensitivity analysis of including individuals with the APOE-4 variant had a minor impact on the results on cognitive decline rates and the onset time of accelerated cognitive decline among minoritized groups, and it did not change the main conclusions regarding average cognitive differences and the evidence of accelerated cognitive decline among all demographic groups.

Much has been written about Medicare expenses in the last phase of life, with estimates ranging from 13% to 25% during the last year, contingent on various methodologies and assumptions (Aldridge & Kelley, 2015; Duncan et al., 2019; Riley & Lubitz, 2010). Given the rising proportion of older adult individuals due to population aging, and particularly the increasing representation of underrepresented populations in population projections, (Colby & Ortman, 2015) healthcare spending has emerged as a critical concern for the sustainability of the healthcare system. Our work shows all respondents undergo cognitive decline and especially during the final phase of life, imposing an additional burden on healthcare systems that may not be adequately prepared to handle the anticipated surge in demand for care and medical expenses. For patients to get goal-concordant care, the Advanced Care Planning (ACP) process should start much earlier. Especially for racial-ethnic minoritized groups, who exhibited lower cognitive levels compared to NHW residents during the early and accelerated phases of cognitive decline. We need to implement culturally respectful systems of care to support older adults from diverse communities age and die at home despite cognitive decline that will happen in the last phase of life.

This article relied on HRS data, a longitudinal nationally representative study that provides objective measures of cognitive function. Our HRS data contained a large sample of repeated measures during 20 years of follow-up that allowed us to build statistically robust trajectories of cognitive function for both women and men and racial-ethnic survivors and decedents. We used a multilevel approach that accounted for within and between individual cognitive variation. To estimate the onset time of accelerated cognitive decline, we utilized a change-point model to investigate the relationship between cognitive decline and the timing of death. Our analysis included the mortality status and restricted the sample to individuals without the APOE-4 variant to account for the influence of mortality on the outcome of decedents and selective survival. The results and conclusions remained unchanged when we tested the performance and generalization ability of our model using a well-established technique in statistics and machine learning for out-of-sample prediction.

### Limitations

There are some limitations in this article that should be noted: 1) since HRS data is reported biennially and we grouped age in 5-year bins to improve the statistical power of our estimates, perhaps resulted in less detailed trajectories of cognitive decline and the onset time of the accelerated decline phase; 2) the sex and race-ethnic differences and age trajectories in cognition were primarily based on word recall and mental status scores; therefore, these findings might not necessarily apply to other cognitive or dementia outcomes, which could exhibit different patterns; 3) although our statistical models were robust and convergence was not an issue, it is worth mentioning that the sample sizes for Hispanic and Black respondents were substantially smaller compared to those for NHW respondents, and this was reflected in the precision of our estimates for underrepresented groups; and 4) although we made efforts to incorporate mortality status into the analysis and conducted sensitivity checks, there may still be persistent effects on sex and race-ethnic disparities due to the competing risk of death and selective survival (Shaw et al., 2021b; Tan et al., 2021).

## CONCLUSION

This paper shows gender and race-ethnic differences in cognitive decline across ages, with women performing better than men in word recall but men better than women in mental status, irrespective of race-ethnicity. In contrast, Black respondents, irrespective of sex, generally demonstrated lower performance in both word recall and mental status compared to Hispanic and White respondents. Moreover, the study presents robust evidence of accelerated cognitive decline, wherein cognition rates decreased at an accelerated pace in the years leading up to death. The onset time, rates of decline, and levels of cognitive function differ not only between men and women but also among different racial-ethnic groups. This research has potential clinical and policy implications. It can help identify and address biases or discriminatory practices, allocate resources effectively to areas or communities in greatest need, and facilitate the development of evidence-based interventions to reduce disparities and improve end-of-life care for individuals of all backgrounds. However, to better understand the underlying causes of these observed differences in cognitive decline before death, further research is warranted.

## Data Availability

All data produced in the present study are available upon reasonable request to the authors

## Notes

### Competing Interest Statement

The authors have declared no competing interest.

### Funding Statement

This work was supported by the National Institutes of Health/National Institute of Aging (P30AG059307) to the Stanford Aging and Ethnogeriatrics (SAGE) Research Center.

### Author Declarations

The study used openly available human data from the Health and Retirement Study that were originally located at: https://hrsdata.isr.umich.edu/data-products/public-survey-data?_gl=1*owsh3n*_ga*ODc3ODY3NjA0LjE3MTk1Mjg3ODk.*_ga_FF28MW3MW2*MTcxOTUyODc4OS4xLjEuMTcxOTUyODc5OC4wLjAuMA..

