## Supplementary Material for "Diverging Patterns of Cognitive Decline by Sex and Race-Ethnicity in Seriously Ill Older Americans"

#### Appendix A. Statistical Analysis

To assess changes in cognitive function and cognitive decline before death across ages, we relied on a two-level mixed-effects change point model (Hall et al., 2000; Karr et al., 2018; Sliwinski et al., 2006; Walter et al., 2016), where the model specification is as follows:

Level 1 model:

$$Y_{it} = \beta_{0i} + \beta_{1i} \cdot min(Age_{it}, DeathAge_i - \tau) + \beta_{2i} \cdot max(0, Age - DeathAge_i + \tau) + \beta_{3i}\boldsymbol{X}_{it} + \varepsilon_{it}. \quad (1)$$

*Age trajectories of cognitive decline*

In our first model, we consider all survivors and decedents, ignoring the potential impact of accelerated cognitive decline; that is, we use the model specification in equation 1 with $\tau = 0$, where the parameter τ represents the onset time of the accelerated phase relative to the time of death (in equation 1, when $\tau = 0$, $min(Age_{it}, DeathAge_i - \tau) = Age_{it}$ and $max(0, Age - DeathAge_i + \tau)$=0). In the resulting model, $Y_{it}$ is the summary cognitive score in HRS of respondent *i* at time *t*, for $i=1,\ldots,T_i$, $Age_{it}$ is the age of respondent *i* at time *t*, and $\boldsymbol{X}_{it}$ represents a vector that includes all covariates described in the previous section (except sex and race-ethnicity), including person-level variables (education, APOE) and those that can change with age for every individual (risk factors and health conditions). We included death status as an explanatory factor in $\boldsymbol{X}_{it}$ to account for the impact of mortality on the outcome of decedents in our model. We included a random within-person error term for person *i* at time *t*, denoted by $\varepsilon_{it}$.

To assess sex and race-ethnic differences in cognition by age, we added a Level 2 model below (equations 2 and 3), which includes the influence of sex and race-ethnicity on individual intercepts and age slopes estimated in the Level 1 model.

Level 2 model:

$$\beta_{0i} = \gamma_{00} + \gamma_{01}Sex_i + \gamma_{02}Race_i + \gamma_{03}Sex_i * Race_i + u_{0i}, \qquad (2)$$

$$\beta_{1i} = \gamma_{10} + \gamma_{11}Sex_i + \gamma_{12}Race_i + \gamma_{13}Sex_i * Race_i + u_{1i}. \qquad (3)$$

The hypothesis of modified sex and race-ethnic differences in age trajectories of cognitive decline is tested by the significance of the random coefficients estimates $\gamma_{j1}$, $\gamma_{j2}$, and $\gamma_{j3}$ (j=0,1) in equations 2-3, which also include between-person random errors captured by the terms $u_{0i}$, and $u_{1i}$.

*Cognitive decline in the last phase of life*

To assess the potential of accelerated cognitive decline, we restricted the sample to all decedent respondents, and we relied on the change point model specification of equation 1, which provides an adequate index of time to death, as opposed to traditional approaches that use the chronological age or time from birth (Sliwinski et al., 2006; Wilson et al., 2020).

Therefore, equation 1 represents two change processes: early phase and accelerated cognitive variation phase. Both effects are captured by estimates from $\beta_{1i}$ and $\beta_{2i}$ ($\hat{\beta}_1$, $\hat{\beta}_2$), where $\hat{\beta}_1$ captures the rate of cognitive change during the early phase, and $\hat{\beta}_2$ captures the rate of change during the accelerated phase. The variable $DeathAge_i$ is the age at death for individual *i*. To estimate $\tau$, we used the profile likelihood method proposed by Hall et al. (2000) (Hall et al., 2000). where we fit several models, one for different values of $\tau$ (ranging between 0 to 15), and select the $\hat{\tau}$ that corresponds to the model producing the highest likelihood.

To estimate the influence of age, and sex and race-ethnic differences in cognitive decline during the early and accelerated phases, we continued using Level 1 and 2 models as specified in equations 1-3 for $\beta_1$ and $\beta_2$, but replacing estimated $\hat{\tau}$ value into the Level 1 equation. The resulting $\hat{\beta}_1$ and $\hat{\beta}_2$ parameters represent the normative age-related process during the early phase and the change during the accelerated phase.

*Sensitivity Analysis: out-of-sample validation*

To assess the robustness and generalization capability of our model, we employed a 10-fold cross-validation approach. In this method, we divided the analytical dataset randomly into ten equal subsets. We replicated the original sex and race-ethnic stratified analyses using the change point model (defined by equations 1-3) to estimate rates of cognition change during the early and accelerated phases. We used previously estimated values for $\hat{\tau}$ corresponding to each subpopulation. Specifically, we used nine of the subsets as a training sample and reserved the remaining subset as the validation or testing set. We repeated this process ten times, altering the training and validation sets each time. The final estimates were calculated as the average of the results obtained from the ten validation sets. We provided confidence intervals for each validation set to assess the consistency of coverage across validation sets, serving as evidence of the stability and generalizability of the model.

### Appendix B. Supplementary Figures and Tables

(a) Total Word Recall

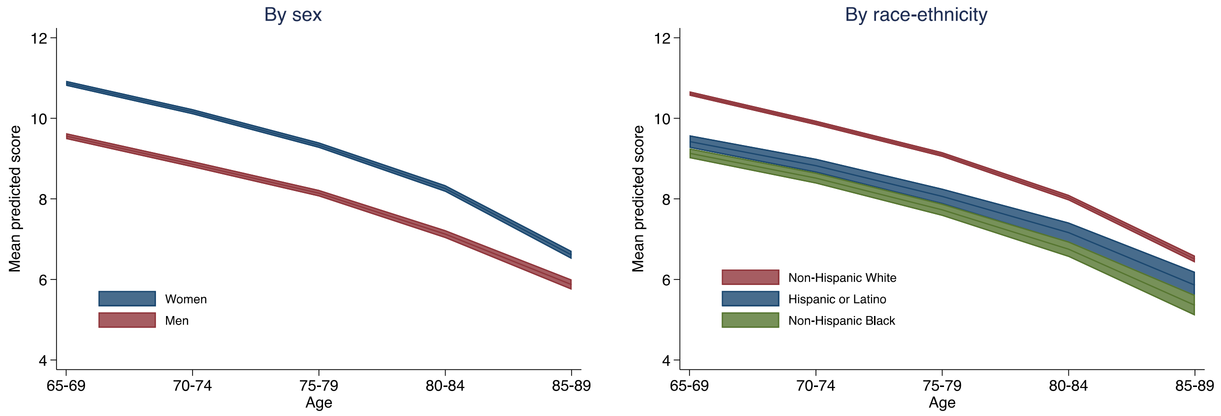


(b) Total Mental Status

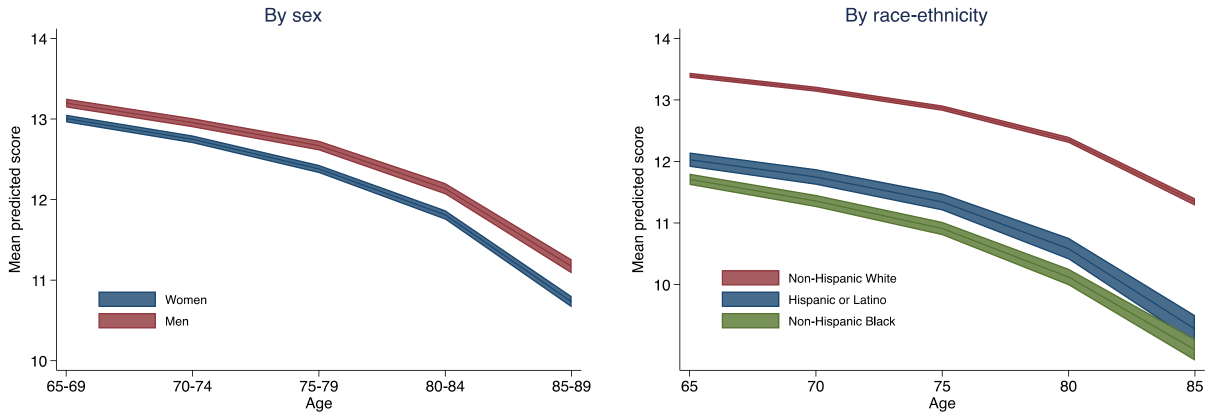


**Figure S1** Predicted summary scores for the total word recall, and total mental status by age, sex, and race-ethnicity.
Note: Age trajectories of cognition were estimated using the model specification of equations 1-3, but with $\tau$=0, and without combining sex and race-ethnic groups.

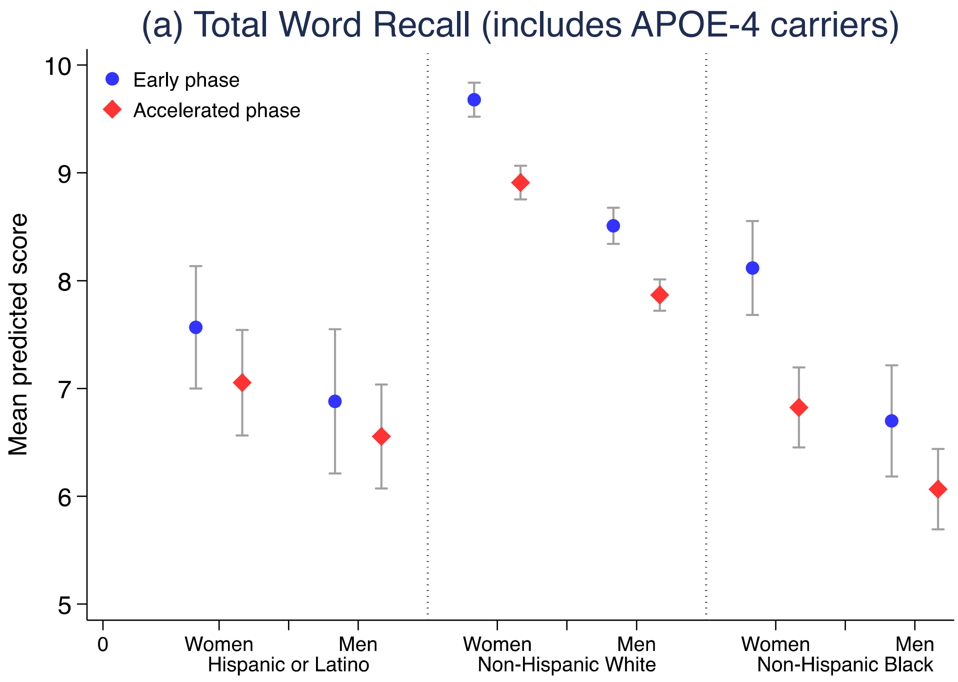


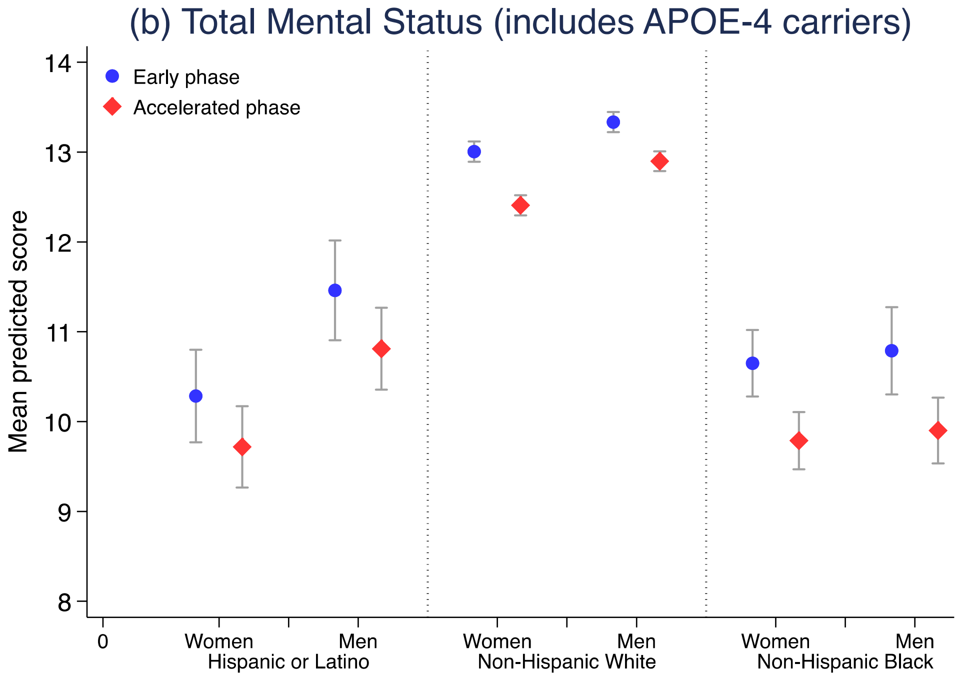


**Figure S2.** Mean predicted word and mental scores at age 75 from HRS decedents by sex and race-ethnicity.

Note: Values predicted cognition were estimated using the change model specification from equations 1-3, while including respondents with the APOE-4 gene variant.

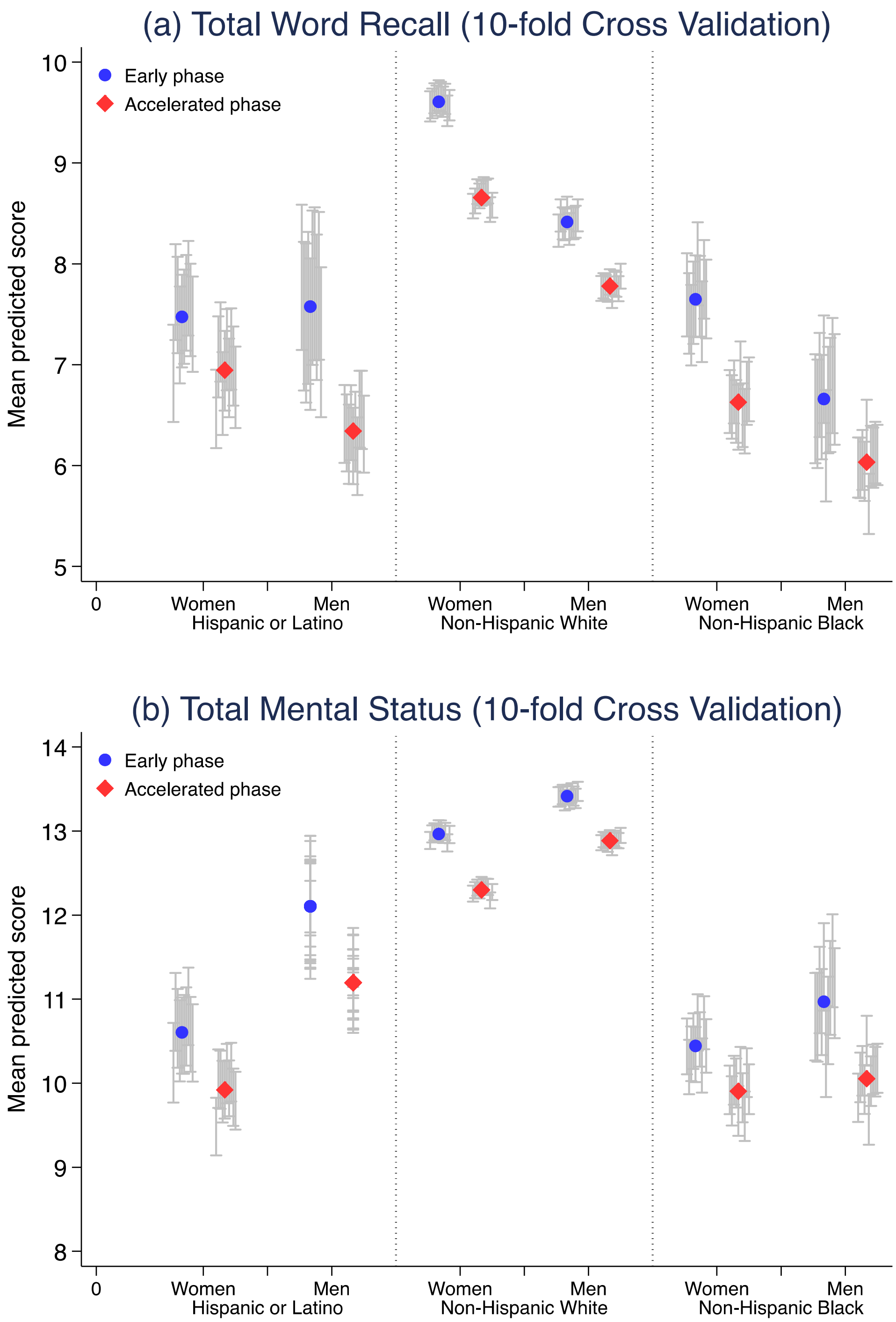


**Figure S3.** Mean predicted word and mental scores at age 75 from HRS decedents by sex and race-ethnicity.

Note: Values predicted cognition were estimated using the change model specification from equations 1-3 as the average of 10 estimates from the ten cross-validation datasets. Error bars represent the 95% CI for each of the ten cross-validation datasets. Consistency of the coverage across datasets provides evidence of the stability and generalizability of the model.

**Table S1.** Point estimates of the onset time of the accelerated phase and rates of early and accelerated cognitive decline across sex and race-ethnic groups

| Cognition Score | Phase 1: Early cognitive decline phase | Phase 2: Accelerated cognitive decline phase | Mean time in years of accelerated cognitive decline (Phase 2) | Annual Rate of Accelerated Decline (ARAD) |
|---|---|---|---|---|
| **Total Word Recall** | | | | |
| Hispanic women | -0.19** (-0.22, -0.17) | -0.27** (-0.31, -0.23) | 10 | -2.6 |
| Hispanic men | -0.16** (-0.19, -0.13) | -0.23** (-0.26, -0.19) | 10 | 0.0 |
| White women | -0.2** (-0.2, -0.19) | -0.37** (-0.38, -0.36) | 10 | -2.2 |
| White men | -0.16** (-0.17, -0.16) | -0.3** (-0.31, -0.29) | 10 | -3.6 |
| Black women | -0.18** (-0.2, -0.16) | -0.32** (-0.35, -0.29) | 10 | 0.0 |
| Black men | -0.14** (-0.16, -0.12) | -0.23** (-0.26, -0.2) | 10 | -2.8 |
| **Total Mental Status** | | | | |
| Hispanic women | -0.13** (-0.16, -0.11) | -0.24** (-0.27, -0.21) | 10 | -2.4 |
| Hispanic men | -0.11** (-0.13, -0.08) | -0.21** (-0.25, -0.18) | 9 | -2.4 |
| White women | -0.08** (-0.09, -0.07) | -0.25** (-0.25, -0.24) | 10 | -2.5 |
| White men | -0.06** (-0.07, -0.06) | -0.21** (-0.22, -0.2) | 9 | -2.3 |
| Black women | -0.12** (-0.13, -0.1) | -0.27** (-0.29, -0.25) | 10 | -2.7 |
| Black men | -0.1** (-0.12, -0.07) | -0.24** (-0.26, -0.21) | 10 | -2.4 |

Note: For this analysis, we replicated results from Table 2 but included respondents with the APOE-4 gene variant. The onset time refers to the accelerated cognitive decline phase relative to the time of death ($\tau$). The ARAD is estimated as the rates of accelerated decline divided by the onset time of accelerated decline. Levels of significance: *(p<0.05), **(p<0.01)

**Table S2.** Mean predicted word and mental scores at ages 65 to 99 from HRS decedents by sex and race-ethnicity: main and sensitivity analyses

| Age Group | Phase | Main Analysis: Excluding APOE-4 carriers | | | Sensitivity Analysis: Including APOE-4 carriers | | | Sensitivity Analysis: 10-fold Cross Validation* | | |
|---|---|---|---|---|---|---|---|---|---|---|
| | | Hispanic o Latino | White | Black | Hispanic o Latino | White | Black | Hispanic o Latino | White | Black |
| **Total Word Recall** | | | | | | | | | | |
| **Women** | | | | | | | | | | |
| 65-69 | Early | 9 (8.5, 9.4) | 10.9 (10.7, 11) | 8.8 (8.4, 9.2) | 9.2 (8.7, 9.7) | 11.2 (11.1, 11.4) | 8.9 (8.5, 9.2) | 8.6 (8.9, 8.5) | 9.4 (11.1, 11) | 11.3 (9, 8.6) |
| 65-69 | Accelerated | 8.3 (7.7, 8.8) | 10.3 (10.1, 10.5) | 8.1 (7.7, 8.4) | 8 (7.3, 8.6) | 10.5 (10.2, 10.7) | 8.1 (7.7, 8.6) | 8.1 (8.3, 7.8) | 8.9 (10.3, 10.1) | 10.5 (8.1, 7.8) |
| 70-74 | Early | 8 (7.6, 8.4) | 10 (9.9, 10.2) | 8 (7.6, 8.4) | 8.1 (7.6, 8.6) | 10.5 (10.3, 10.6) | 8.6 (8.2, 9) | 7.8 (8.1, 7.7) | 8.6 (10.3, 10.1) | 10.4 (8.4, 8) |
| 70-74 | Accelerated | 7.8 (7.4, 8.3) | 9.5 (9.3, 9.6) | 7.3 (7, 7.6) | 7.3 (6.7, 7.8) | 9.7 (9.6, 9.9) | 7.7 (7.3, 8) | 7.4 (7.8, 7.3) | 8.3 (9.5, 9.4) | 9.7 (7.4, 7) |
| 75-79 | Early | 7.3 (6.9, 7.8) | 9.3 (9.2, 9.5) | 7.4 (7, 7.8) | 7.6 (7, 8.1) | 9.7 (9.5, 9.8) | 8.1 (7.7, 8.6) | 6.9 (7.5, 7) | 7.9 (9.6, 9.5) | 9.8 (7.6, 7.2) |
| 75-79 | Accelerated | 6.9 (6.5, 7.3) | 8.6 (8.4, 8.7) | 6.5 (6.2, 6.8) | 7.1 (6.6, 7.5) | 8.9 (8.8, 9.1) | 6.8 (6.5, 7.2) | 6.6 (6.9, 6.5) | 7.3 (8.7, 8.5) | 8.8 (6.6, 6.3) |
| 80-84 | Early | 6.2 (5.6, 6.8) | 8.5 (8.3, 8.6) | 6.6 (6.1, 7.1) | 6.5 (5.8, 7.3) | 9 (8.8, 9.2) | 7.4 (6.8, 8) | 5.7 (6.2, 5.6) | 6.9 (8.8, 8.6) | 9 (7, 6.4) |
| 80-84 | Accelerated | 5.9 (5.5, 6.3) | 7.5 (7.4, 7.6) | 5.7 (5.4, 6) | 6.1 (5.6, 6.7) | 7.8 (7.7, 8) | 6.1 (5.7, 6.6) | 5.7 (6.1, 5.6) | 6.5 (7.7, 7.6) | 7.8 (6, 5.7) |
| 85-99 | Early | 6.2 (5.4, 7) | 7.6 (7.2, 7.9) | 6 (5.1, 6.8) | 6.1 (4.9, 7.3) | 8.1 (7.8, 8.5) | 6.6 (5.5, 7.8) | 6 (6.5, 5.6) | 7.4 (7.8, 7.4) | 8.2 (6.9, 5.9) |
| 85-99 | Accelerated | 4.8 (4.4, 5.3) | 6.1 (5.9, 6.2) | 4.6 (4.3, 4.9) | 5 (4.4, 5.6) | 6.5 (6.3, 6.6) | 4.9 (4.5, 5.4) | 4.7 (5.1, 4.7) | 5.5 (6.4, 6.3) | 6.5 (5.1, 4.8) |
| **Men** | | | | | | | | | | |
| 65-69 | Early | 7.9 (7.5, 8.4) | 9.6 (9.5, 9.8) | 7.5 (7.2, 7.9) | 8.5 (8, 9) | 9.9 (9.7, 10) | 7.7 (7.3, 8.1) | 8.6 (8.4, 7.9) | 8.9 (9.7, 9.6) | 9.9 (7.7, 7.3) |
| 65-69 | Accelerated | 7.3 (6.8, 7.7) | 9.2 (9.1, 9.4) | 7.2 (6.9, 7.6) | 7.5 (6.9, 8.1) | 9.5 (9.3, 9.7) | 6.9 (6.5, 7.4) | 8.1 (7.2, 6.7) | 7.7 (9.3, 9.1) | 9.4 (7.2, 6.9) |
| 70-74 | Early | 7.3 (6.9, 7.8) | 8.9 (8.8, 9.1) | 7 (6.6, 7.4) | 7.6 (7.1, 8.1) | 9.2 (9, 9.3) | 7.1 (6.7, 7.5) | 7.8 (7.9, 7.4) | 8.5 (9.1, 8.9) | 9.2 (7.1, 6.6) |

| | | | | | | | | | | |
|---|---|---|---|---|---|---|---|---|---|---|
| 70-74 | Accelerated | 7 (6.6, 7.4) | 8.4 (8.2, 8.5) | 6.3 (6, 6.6) | 7.6 (7.1, 8.1) | 8.6 (8.4, 8.7) | 6.6 (6.3, 7) | 7.4 (7, 6.6) | 7.5 (8.4, 8.2) | 8.5 (6.4, 6.1) |
| 75-79 | Early | 6.6 (6, 7.3) | 8.2 (8.1, 8.4) | 6.5 (6, 7) | 6.9 (6.2, 7.5) | 8.5 (8.3, 8.7) | 6.7 (6.2, 7.2) | 6.9 (7.6, 6.8) | 8.3 (8.4, 8.3) | 8.6 (6.7, 6.1) |
| 75-79 | Accelerated | 6.1 (5.7, 6.5) | 7.7 (7.6, 7.9) | 5.8 (5.5, 6.1) | 6.6 (6.1, 7) | 7.9 (7.7, 8) | 6.1 (5.7, 6.4) | 6.6 (6.3, 6) | 6.7 (7.8, 7.7) | 7.9 (6, 5.7) |
| 80-84 | Early | 5.5 (4.7, 6.4) | 7.6 (7.4, 7.8) | 5.6 (4.9, 6.3) | 6.3 (5.3, 7.2) | 7.8 (7.6, 8) | 6.4 (5.6, 7.2) | 5.7 (6.5, 5.6) | 7.5 (7.8, 7.6) | 8 (5.3, 4.5) |
| 80-84 | Accelerated | 5.2 (4.8, 5.7) | 6.8 (6.6, 6.9) | 5.4 (5.1, 5.8) | 5.8 (5.2, 6.3) | 7 (6.8, 7.1) | 5.5 (5.1, 5.9) | 5.7 (5.5, 5) | 6 (6.9, 6.7) | 7 (5.7, 5.3) |
| 85-99 | Early | 5.1 (3.5, 6.8) | 7 (6.7, 7.4) | 5.6 (4.4, 6.8) | 5.7 (3.6, 7.7) | 7.2 (6.7, 7.7) | 6.2 (4.6, 7.8) | 6 (5.2, 3.2) | 7.2 (7.4, 7) | 7.9 (5.9, 4.5) |
| 85-99 | Accelerated | 4.5 (3.9, 5.1) | 5.8 (5.6, 5.9) | 4.6 (4.1, 5) | 4.7 (4, 5.4) | 5.9 (5.7, 6) | 5 (4.4, 5.5) | 4.7 (5, 4.4) | 5.5 (6, 5.9) | 6.1 (4.8, 4.4) |
| **Total Mental Status** | | | | | | | | | | |
| **Women** | | | | | | | | | | |
| 65-69 | Early | 11 (10.7, 11.4) | 13.2 (13.1, 13.3) | 10.7 (10.4, 11) | 11.1 (10.6, 11.5) | 13.5 (13.3, 13.6) | 11 (10.7, 11.3) | 10.4 (10.7, 10.3) | 11.2 (13.2, 13) | 13.3 (10.6, 10.2) |
| 65-69 | Accelerated | 10.6 (10.1, 11) | 12.8 (12.7, 13) | 10.3 (10, 10.7) | 10.5 (9.9, 11.1) | 13.1 (12.9, 13.2) | 10.5 (10.1, 10.8) | 10.3 (10.6, 10.2) | 11.1 (12.8, 12.6) | 12.9 (10.3, 10) |
| 70-74 | Early | 11 (10.6, 11.4) | 13 (12.9, 13.1) | 10.4 (10.2, 10.7) | 11 (10.6, 11.5) | 13.2 (13.1, 13.3) | 10.8 (10.4, 11.1) | 10.6 (11, 10.6) | 11.5 (13, 12.9) | 13.2 (10.7, 10.3) |
| 70-74 | Accelerated | 10.3 (9.9, 10.7) | 12.6 (12.4, 12.7) | 9.8 (9.6, 10.1) | 10.3 (9.9, 10.8) | 12.7 (12.6, 12.9) | 10.1 (9.8, 10.4) | 9.9 (10.2, 9.8) | 10.7 (12.6, 12.4) | 12.7 (9.6, 9.3) |
| 75-79 | Early | 10.5 (10, 10.9) | 12.8 (12.7, 12.9) | 10.2 (9.9, 10.5) | 10.3 (9.8, 10.8) | 13 (12.9, 13.1) | 10.6 (10.3, 11) | 10.2 (10.6, 10.1) | 11.1 (13, 12.9) | 13.1 (10.4, 10.1) |
| 75-79 | Accelerated | 9.8 (9.5, 10.2) | 12.3 (12.2, 12.4) | 9.7 (9.4, 10) | 9.7 (9.3, 10.2) | 12.4 (12.3, 12.5) | 9.8 (9.5, 10.1) | 9.5 (9.9, 9.6) | 10.3 (12.3, 12.2) | 12.4 (9.9, 9.6) |
| 80-84 | Early | 10 (9.5, 10.6) | 12.5 (12.4, 12.6) | 9.8 (9.5, 10.2) | 10.1 (9.5, 10.8) | 12.8 (12.6, 12.9) | 10.2 (9.7, 10.7) | 9.7 (10.3, 9.7) | 11 (12.7, 12.6) | 12.8 (10.3, 9.9) |
| 80-84 | Accelerated | 9.1 (8.8, 9.5) | 11.7 (11.6, 11.8) | 8.8 (8.5, 9.1) | 9.1 (8.6, 9.6) | 11.9 (11.8, 12) | 9 (8.7, 9.4) | 9 (9.3, 8.9) | 9.7 (11.8, 11.7) | 11.9 (9.3, 9) |
| 85-99 | Early | 9.2 (8.4, 9.9) | 12.2 (12, 12.4) | 9 (8.5, 9.6) | 9.1 (8.2, 10.1) | 12.5 (12.2, 12.7) | 9.3 (8.5, 10.2) | 9.8 (9.7, 8.7) | 10.7 (12.3, 12.1) | 12.6 (9.8, 9.1) |
| 85-99 | Accelerated | 8 (7.6, 8.4) | 10.8 (10.7, 10.9) | 7.8 (7.5, 8.1) | 7.8 (7.3, 8.3) | 11 (10.9, 11.1) | 8.2 (7.8, 8.6) | 8.1 (8.5, 8.1) | 8.8 (11.1, 11) | 11.2 (8.4, 8.2) |

| Men | | | | | | | | | | |
|---|---|---|---|---|---|---|---|---|---|---|
| 65-69 | Early | 11.9 (11.5, 12.3) | 13.6 (13.5, 13.7) | 11.2 (10.9, 11.5) | 11.9 (11.5, 12.4) | 13.7 (13.6, 13.8) | 11.2 (10.9, 11.6) | 10.4 (11.9, 11.4) | 12.3 (13.4, 13.3) | 13.5 (11.3, 10.9) |
| 65-69 | Accelerated | 11.9 (11.4, 12.3) | 13.3 (13.2, 13.4) | 10.8 (10.5, 11.2) | 11.8 (11.2, 12.4) | 13.4 (13.3, 13.6) | 11 (10.6, 11.4) | 10.3 (11.6, 11.2) | 12.1 (13.2, 13.1) | 13.3 (10.6, 10.2) |
| 70-74 | Early | 12 (11.6, 12.4) | 13.5 (13.4, 13.6) | 10.7 (10.4, 11.1) | 11.7 (11.2, 12.2) | 13.5 (13.4, 13.6) | 11 (10.6, 11.4) | 10.6 (12.3, 11.8) | 12.8 (13.4, 13.3) | 13.5 (10.9, 10.5) |
| 70-74 | Accelerated | 11.3 (10.9, 11.6) | 13.1 (13, 13.2) | 10.4 (10.1, 10.7) | 11.5 (11, 12) | 13.1 (13, 13.2) | 10.5 (10.1, 10.8) | 9.9 (11.2, 10.8) | 11.6 (13, 12.9) | 13.1 (10.5, 10.2) |
| 75-79 | Early | 11.6 (11.1, 12.1) | 13.3 (13.2, 13.4) | 10.7 (10.3, 11.1) | 11.5 (10.9, 12) | 13.3 (13.2, 13.4) | 10.8 (10.3, 11.3) | 10.2 (12.1, 11.5) | 12.7 (13.4, 13.3) | 13.5 (11, 10.4) |
| 75-79 | Accelerated | 10.9 (10.6, 11.3) | 12.9 (12.8, 13) | 9.8 (9.5, 10.1) | 10.8 (10.4, 11.3) | 12.9 (12.8, 13) | 9.9 (9.5, 10.3) | 9.5 (11.2, 10.8) | 11.6 (12.9, 12.8) | 13 (10.1, 9.8) |
| 80-84 | Early | 11.6 (10.9, 12.2) | 13 (12.9, 13.2) | 10.5 (9.9, 11.1) | 10.9 (10.2, 11.7) | 13.1 (12.9, 13.2) | 10.4 (9.7, 11.1) | 9.7 (11.8, 11) | 12.6 (13.3, 13.1) | 13.4 (10.6, 9.8) |
| 80-84 | Accelerated | 10.2 (9.8, 10.6) | 12.4 (12.3, 12.5) | 9.6 (9.3, 9.9) | 10.2 (9.7, 10.7) | 12.3 (12.2, 12.4) | 9.4 (9, 9.8) | 9 (10.4, 10) | 10.8 (12.6, 12.5) | 12.6 (10.1, 9.7) |
| 85-99 | Early | 10.6 (9.6, 11.7) | 12.8 (12.6, 13.1) | 10.4 (9.3, 11.4) | 10.3 (9.1, 11.5) | 12.9 (12.6, 13.2) | 10 (8.6, 11.4) | 9.8 (10, 8.5) | 11.5 (13.2, 12.9) | 13.5 (10.9, 9.5) |
| 85-99 | Accelerated | 9.5 (9, 10) | 11.5 (11.4, 11.7) | 8.4 (8, 8.8) | 9.1 (8.5, 9.8) | 11.6 (11.4, 11.7) | 8.3 (7.8, 8.8) | 8.1 (9.8, 9.3) | 10.4 (11.9, 11.8) | 12 (9.1, 8.7) |

*Notes*: Values predicted cognition were estimated using the change model specification from equations 1-3, for the main and sensitivity models as described above in Appendix A.

* Confidence intervals are computed as the average of those resulting from the 10-fold cross-validation results.
